# Variations in determining actual orientations of segmented deep brain stimulation leads using the manually refined DiODe algorithm: a retrospective study across different lead designs and medical institutions

**DOI:** 10.1101/2022.12.22.22283840

**Authors:** Kaylee R. Henry, Milina Miulli, Noa Nuzov, Mark J. Nolt, Joshua Rosenow, Behzad Elahi, Julie G. Pilitsis, Laleh Golestanirad

**Affiliations:** Department of Biomedical Engineering, Northwestern University, Evanston, IL, USA; Department of Neuroscience and Department of Global Health Studies, Northwestern University, Evanston, IL, USA; Department of Neurosurgery, Feinberg School of Medicine, Northwestern University, Chicago, IL, USA; Department of Physical Therapy and Human Movement Sciences, Northwestern University, Chicago, IL, USA; Department of Neurosciences & Experimental Therapeutics, Albany Medical College, Albany, NY, USA; Department of Radiology, Northwestern University, Chicago, IL, USA

**Keywords:** deep brain stimulation (DBS), directional orientation detection (DiODe), directional leads, Lead-DBS, user agreement, manual refinement

## Abstract

**Purpose:** Directional deep brain stimulation (DBS) leads have become widely used in the past decade. Understanding the asymmetric stimulation provided by directional leads requires precise knowledge of the exact orientation of the lead in respect to its anatomical target. Recently, the DiODe algorithm was developed to automatically determine the orientation angle of leads from the artifact on postoperative computed tomography (CT) images. However, DiODe results are user-dependent. This study analyzed the significance of lead rotation as well as the user agreement of DiODe calculations across the two most common DBS systems and two independent medical institutions.

**Methods:** Data from 104 patients who underwent an anterior-facing unilateral/bilateral directional DBS implantation at either Northwestern Memorial Hospital (NMH) or Albany Medical Center (AMC) were retrospectively analyzed. Actual orientations of the implanted leads were independently calculated by three individual users using the DiODe algorithm in Lead-DBS and patients’ postoperative CT images. Deviation from the intended orientation and user agreement were assessed.

**Results:** All leads significantly deviated from the intended 0° orientation (p<0.001), regardless of DBS lead design (p<0.05) or institution (p<0.05). However, a bias of the implantation towards a single direction was seen for the Boston Scientific leads (p=0.014 at NMH, p=0.029 at AMC). A difference of 10° between at least two users occurred in 28% (NMH) and 39% (AMC) of all Boston Scientific and 53% (AMC) and 76% (NMH) of all St. Jude leads.

**Conclusion:** Our results show that there is a significant lead rotation from the intended surgical orientation across both DBS systems and both medical institutions, however, a bias towards a single direction was only seen in Boston Scientific leads. Additionally, these results raise questions into the user error that occurs when manually refining the orientation angles calculated with DiODe.

## Introduction

Deep brain stimulation (DBS) as a therapeutic approach to a variety of neurological and psychiatric disorders has heavily relied on neuroimaging for precise electrode placement [1,2]. Recently, the application of neuroimaging for patient-specific DBS programming has also gained traction, both due to concerted efforts to make safe post-operative magnetic resonance imaging (MRI) accessible to DBS patients (1–6), and thanks to studies that demonstrate added benefits of MRI-based DBS management (7–9). This trend is likely to continue as the feasibility and benefits of high and ultra-high field MRI for DBS is explored (10–12).

Historically, the standard DBS lead design consisted of four cylindrical, omnidirectional ring contacts that provided axially symmetrical stimulation of the surrounding neural tissue. Programming of such devices largely relied on clinical trial and error, where individual contacts were activated to find thresholds for therapeutic effects and adverse effects (13). More recently, directional leads have been increasingly implanted, as they allow for asymmetric stimulation of the surrounding neural tissue to reduce therapeutic amplitudes (14) and increase side effect thresholds (15). The two most commonly used directional lead models, namely, Boston Scientific Cartesia™ (Marlborough, MA, USA) and Abbott St. Jude Medical 6172 Directional leads (Chicago, IL, USA) have similar designs: each consisting of two cylindrical ring contacts at the most proximal and distal ends of the lead and two rows of three-segmented contacts in between (shown in Fig. 1A).

**Fig. 1.**
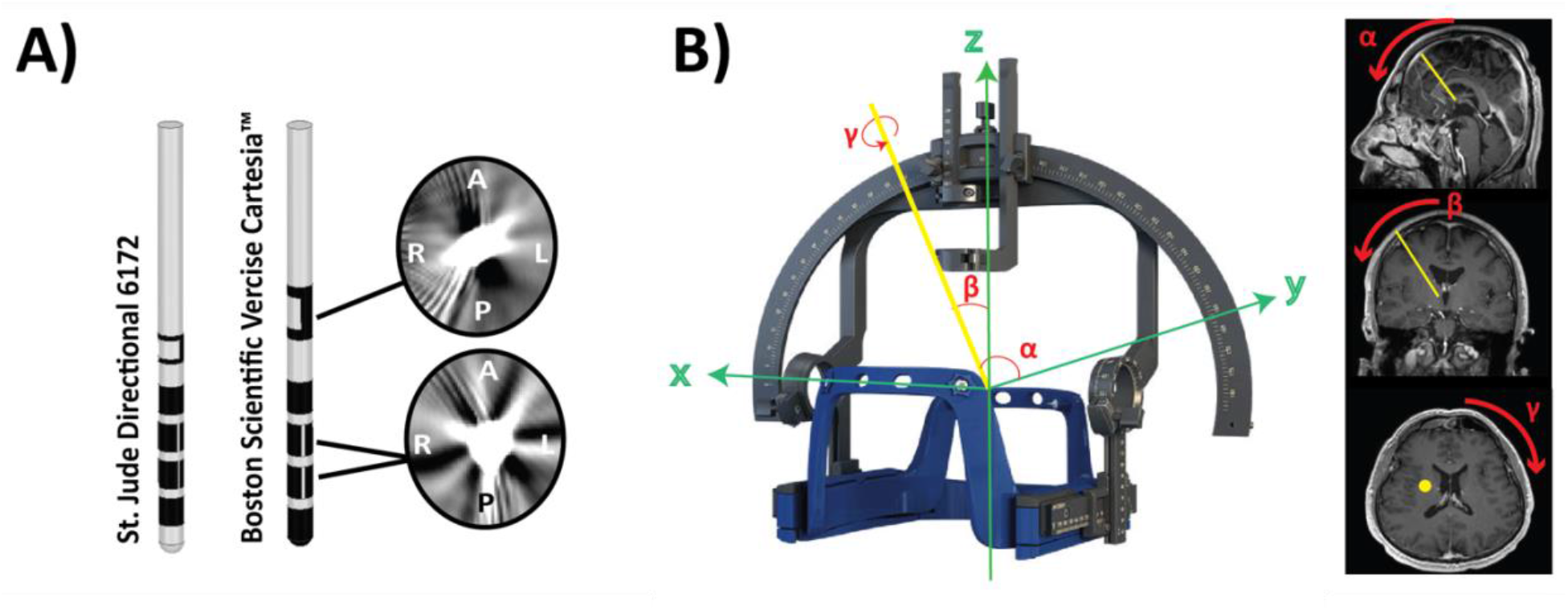
(A) Illustration of the Boston Scientific Cartesia™ and Abbott St. Jude Medical Directional leads and the unique CT artifacts that occur at the levels of the marker and segmented contacts. (B) A stereotactic frame showing the pitch (α), yaw (β), and orientation (γ) angles relative to a DBS lead. Pitch is the rotation around the x-axis, yaw is the rotation around the y-axis, and the orientation angle is lead rotation around the lead’s axis.

Eight-contact directional leads offer many advantages compared to the traditional four-contact omnidirectional leads, including the ability of current steering in the case of a malpositioned lead (16,17) or targeting a brain region that is difficult to stimulate due to the patient’s specific anatomy (18,19), as well as increasing side-effect thresholds, and decreasing efficacy thresholds (14,15). With these advantages, though, comes an unprecedented number of degrees of freedom in DBS programming. It is crucial to accurately understand the location and orientation of the directional DBS leads in regard to surrounding anatomy when clinically interpreting DBS response and avoiding stimulation of specific brain regions that can cause severe psychiatric and motor side effects (20). This is particularly important for the reliable application of recent image-guided programming approaches that are based on computational models of current diffusion within patient-specific anatomy (21–23).

During the implantation of DBS leads, surgeons use a stereotactic frame to establish the exact pitch, yaw, and roll angle for each lead to ensure accurate placement within the target brain region (shown in Fig. 1B). For directional DBS systems, the orientation angle of the lead adds a new degree of freedom which is hard to accurately establish using currently available surgical apparatuses. Although surgeons try to visually control for this angle by attempting to face the lead’s marker toward a specific direction, the accuracy of the achieved angles are not well established.

Recently, the Directional Orientation Detection (DiODe) algorithm was developed to automatically determine the orientation angle of leads from artifacts seen in postoperative computed tomography (CT) images (24,25). To achieve this, DiODe calculates the exact orientation angle of the lead by analyzing unique metal artifacts that occur in CT images at levels of the marker and two-segmented electrodes (shown in Fig. 1A). In addition to being validated through multiple phantom studies, DiODe has also produced highly correlated results with orientations determined from rotation fluoroscopy images, making the algorithm a reliable method of lead rotation analysis in DBS research (26–28).

Using the first version of the DiODe algorithm implemented in version 2.5 of the Lead-DBS software (29), three independent studies reported large deviations of up to 90° between the intended and actual lead orientations in patients (26,27,30). Notably, however, the original DiODe algorithm needed manual interventions by selecting CT slices on which the artifacts were most visible, making its output user-dependent. Also, due to the 180° symmetry of the artifacts, it offered two possible inverse solutions for the lead orientation without being able to distinguish between them. Recently, DiODe v2 has been introduced as an open-source algorithm (https://github.com/Till-Dembek/DiODe_Standalone, accessed on 10 October 2021) which considers the center of mass of the lead artifact to offer a fully automated workflow for selecting the optimal artifact slice and resolves the ambiguity of the artifact symmetry (31). To our knowledge, there have yet to be any studies assessing user agreement in the application of the DiODe v2 algorithm. Moreover, there has yet to be a study analyzing the deviation between the intended and actual orientation of leads across the two most common DBS systems, namely, Boston Scientific Vercise and St. Jude Medical Infinity, or across medical institutions. In this paper, we examined deviations across DBS systems and institutions while also assessing the degree of user agreement.

## Methods

### Ethics

Institutional Review Boards of Northwestern University and Albany Medical Center approved the use of existing imaging data for the purpose of the analysis. Due to the study’s retrospective nature, informed consent was not required from patients. All images were gathered as part of routine clinical care.

### Patient demographics

We retrospectively analyzed all patients (n=139 patients, 220 leads) who underwent unilateral or bilateral directional DBS implantation surgery at Northwestern Memorial Hospital and Albany Medical Center between the years of 2018 and 2022 and for whom the intended orientation of the lead was anterior-posterior. All patients examined had either the Boston Scientific Vercise or Abbott St. Jude Medical Infinity DBS system.

In total, 220 leads were implanted across both institutions. Fifty leads were removed from the analysis due to having a polar angle greater than 40°, which has been reported to result in inaccurate calculations within the DiODe algorithm (25). Of the 170 leads used in the analysis, 142 (83.5%) had been implanted into the subthalamic nucleus (STN), 18 (10.6%) in the ventral intermediate nucleus of the thalamus (VIM), 6 (3.5%) in the globus pallidus internus (GPi), and 1 (0.6%) in the ventral capsule/ventral striatum (VC/VS). Overall, 75% patients were male and the average age was 65±10 years. All leads were implanted facing anterior. Demographic details for the patients are displayed in Table 1.

**Table 1.** Albany Medical Center and Northwestern Memorial Hospital patient demographics.

| Albany Medical Center<br>(n=38 patients) | Northwestern Memorial Hospital<br>(n=66 patients) |
| --- | --- |
| **DBS System:** | **DBS System:** |
| 11 Boston Scientific Vercise | 46 Boston Scientific Vercise |
| 27 Abbott St. Jude Medical<br>Infinity | 20 Abbott St. Jude Medical<br>Infinity |
| **Sex:** | **Sex:** |
| 30 Male | 48 Male |
| 8 Female | 18 Female |
| **Target Brain Region:** | **Target Brain Region:** |
| 5 VIM | 9 VIM |
| 3 GPi | 1 GPi |
| 29 STN | 56 STN |
| 1 VC/VS |  |
| **Laterality:** | **Laterality:** |
| 14 Unilateral | 21 Unilateral |
| 24 Bilateral | 44 Bilateral |

### Surgical procedure

All DBS implantations were performed by J.R. at Northwestern Memorial Hospital (NMH) or J.P. at Albany Medical Center (AMC). Prior to the surgery, Brainlab Elements (Brainlab, Munich, Germany) planning software was used to plan the patient’s lead trajectories using preoperative magnetic resonance images (MRI). Targets were selected using a combination of stereotactic atlas-based coordinates and direct target visualization on MRI. During the surgery, target and entry point coordinates for the pre-planned lead trajectory were set on a Leksell model G stereotactic frame (Elekta, Stockholm, Sweden). A burr hole was drilled at that location, and microelectrode recording and stimulation was conducted to locate the target brain structure and test a simple range of cathodic stimulations (1-3mA, pulse width of 60µs, pulse frequency of 130Hz) (Alpha Omega, Nazareth, Israel at NMH and FHC, Maine, USA at AMC). Optimal trajectories were those with greater than 4 mm of target single unit recordings where there were appropriate neuronal responses to passive limb movements coupled with the presence of stimulation-associated benefits and a lack of significant stimulation-associated side effects. DBS electrodes were measured for implant using the standard measuring bracket and rotated to the appropriate orientation by hand prior to insertion into the brain. To aid in initial orientation alignment, the microdrive adaptor for the Leksell frame was placed in a parallel orientation to the frame arc to maintain anteroposterior alignment to the intercommissural line and reduce rotation out of that plane that could also lead to lead rotation. Leads were placed under intermittent fluoroscopy after alignment of the fluoroscope to the frame reticles. The directional lead marker was visualized on fluoroscopy after implant for concordance with intended orientation.

### Image acquisition

All patients received their initial postoperative CT imaging within 24 hours of the implantation surgery as part of the routine clinical protocol at both Northwestern Memorial Hospital and Albany Medical Center.

### Determining the actual orientation of the DBS leads

The original user-supervised DiODe algorithm, which was implemented in IDL (Exelis Visual Information Solution, USA), has been extensively validated in both geometrical and anthropomorphic phantoms (24,25). An adaptation of DiODe implemented in MATLAB version 2021b (MathWorks, USA) is now available and fully integrated into the open-source Lead-DBS toolbox, version 2.6 (www.lead-dbs.org) (29,32).

DBS lead localization was independently completed by three users (K.H., M.M., and N.N.) using the Lead-DBS manual pre-reconstruction toolbox. Each user then ran DiODe v2 within Lead-DBS, while utilizing the manual refinement option, to calculate the actual orientation angles of each DBS lead detected in the postoperative CT images. The actual orientation angles were recorded for statistical testing. If all users had a strong agreement for a lead (e.g., all calculated orientations fell within 10° of each other), the average orientation was used for the statistical analysis. Cases where the three users had varied results for the actual orientation of a lead (e.g., greater than a 10° difference between at least two users) were noted and blindly re-analyzed by all three users. If there was a strong agreement between at least two users after rerunning DiODe, the orientations of the two users in agreement were averaged. However, if there was still a greater than 10° variation between all users’ actual orientations for that lead, the lead was discarded.

### Statistical analysis

All statistical tests were conducted in RStudio version 1.4.1103 (Boston, MA, USA) with an α of less than 0.05 considered significant. Prior to analyzing the orientation angles, a two one-sided test (TOST), a test of equivalence, was applied to determine if the actual orientation angles estimated by each user (K.H., M.M., and N.N.) were equivalent within a predefined range of ±30° (Fig. 2). The 30° angle was selected because it is the maximum rotation that can occur before the overlapping region of electrode contacts is less than the non-overlapping regions. To determine if the deviations between the intended and actual lead orientations were significant, a two-tailed Wilcoxon Signed-Rank test was applied. Bias was determined with a one-sample, one-tailed Wilcoxon Signed-Rank test.

**Fig. 2.**
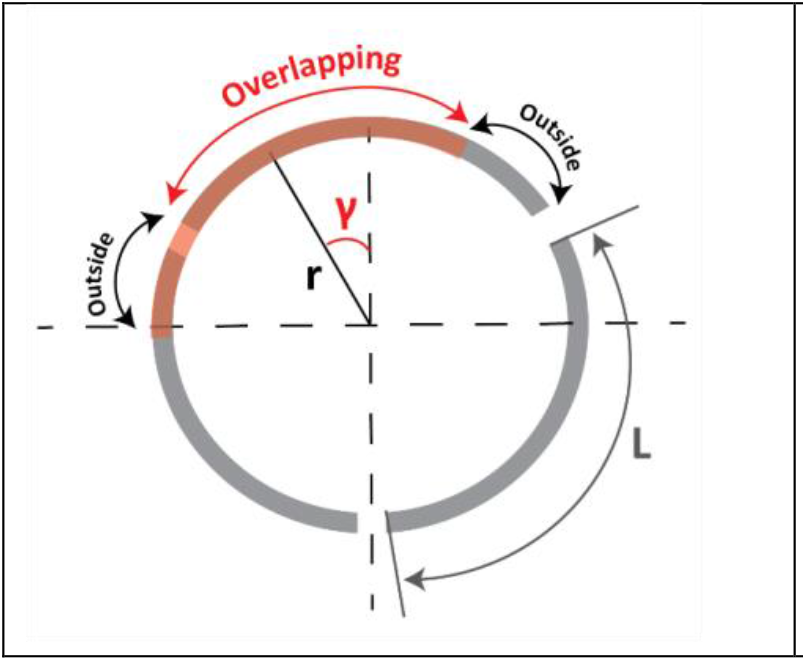
Rotating a contact from its original position (gray) to a new position (red) based upon its orientation (γ). The overlapping region is the arc length shared by the reoriented and original contact, where the outside regions are where only the reoriented or the original contact are located. The angle that leads to the length of overlapping region to be equal to the outside region is *L*/3*r*, where *L* is the arc length of the contact and *r* is the radius of the lead. For directional leads, *L*=1mm and *r*=0.65mm, leading to a γ=30°.

## Results

### User agreement

There was a difference greater than 10° between at least two users in 23/82 (28%) of the Boston Scientific and 19/25 (76%) of the Abbott leads in the Northwestern Memorial Hospital patients. Similarly, we saw this difference between at least two users in 7/18 (38.9%) of the Boston Scientific and 24/45 (53.3%) of the Abbott leads in the Albany Medical Center patients.

There was a greater than 20° difference between at least two users in 13/82 (15.9%) of the Boston Scientific and 15/25 (60%) of the Abbott leads in the Northwestern Memorial Hospital patients. We saw this difference between at least two users in 4/18 (22.2%) of the Boston Scientific and 20/45 (44.4%) of the Abbott leads in the Albany Medical Center patients.

There was a greater than 30° difference between at least two users in 9/82 (10.9%) of the Boston Scientific and 11/25 (44%) of the Abbott leads in the Northwestern Memorial Hospital patients. We saw this difference between at least two users in 3/18 (13.7%) of the Boston Scientific and 17/45 (37.8%) of the Abbott leads in the Albany Medical Center patients.

Even after the blind reanalysis of the leads that had a difference greater than 10° between at least two users, 7 leads (4 leads from NMH: 1 Boston Scientific and 3 St. Jude; 3 leads from AMC: all St. Jude) still did not have agreeing orientations and were discarded.

The median deviation between users K.H. and M.M. was 2° (IQR=4°); between K.H. and N.N. was 2° (IQR=3°); between M.M. and N.N. was 2° (IQR=3°). The TOST confirmed that user estimations of actual angles from DiODe were statistically equivalent within a range of ±30° (p<0.001 for each combination of two users; CI_KH,MM_ = [-8.97°, 7.06°]; CI_KH,NN_ = [-6.56°, 9.84°]; CI_MM,NN_ = [-5.66°, 10.86°]). Each user’s orientation angles as well as the user agreement percentages are illustrated in Figure 3.

**Fig. 3.**
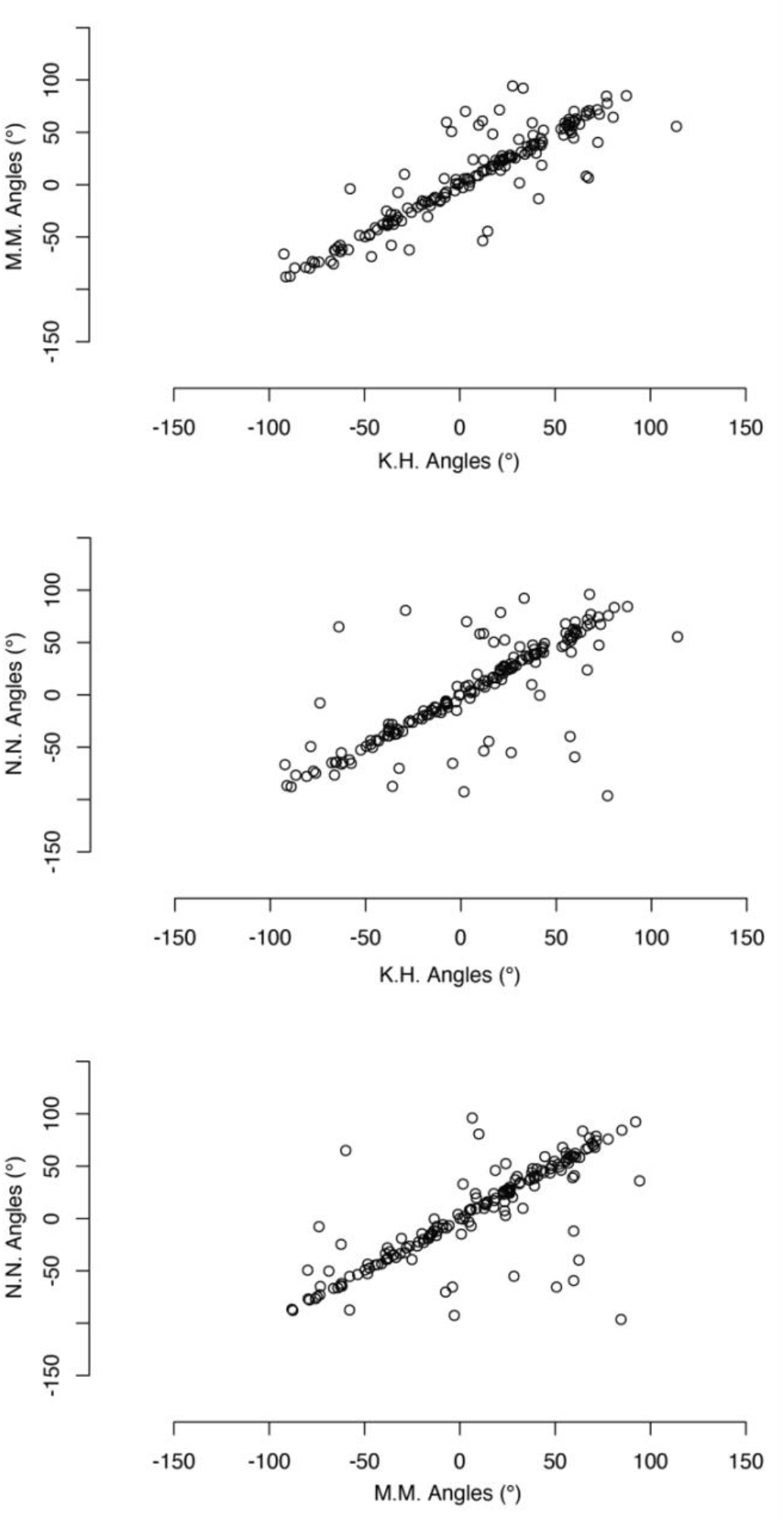
Orientation angles calculated with DiODe across three different users. User agreement was calculated between each combination of users.

### Deviation from intended orientation: all leads

The actual orientations in comparison to the intended orientation are listed in Table 2. Orientation angles were transformed into dimensionless values to ensure that the right and left orientations would not average out to 0°.

**Table 2.**
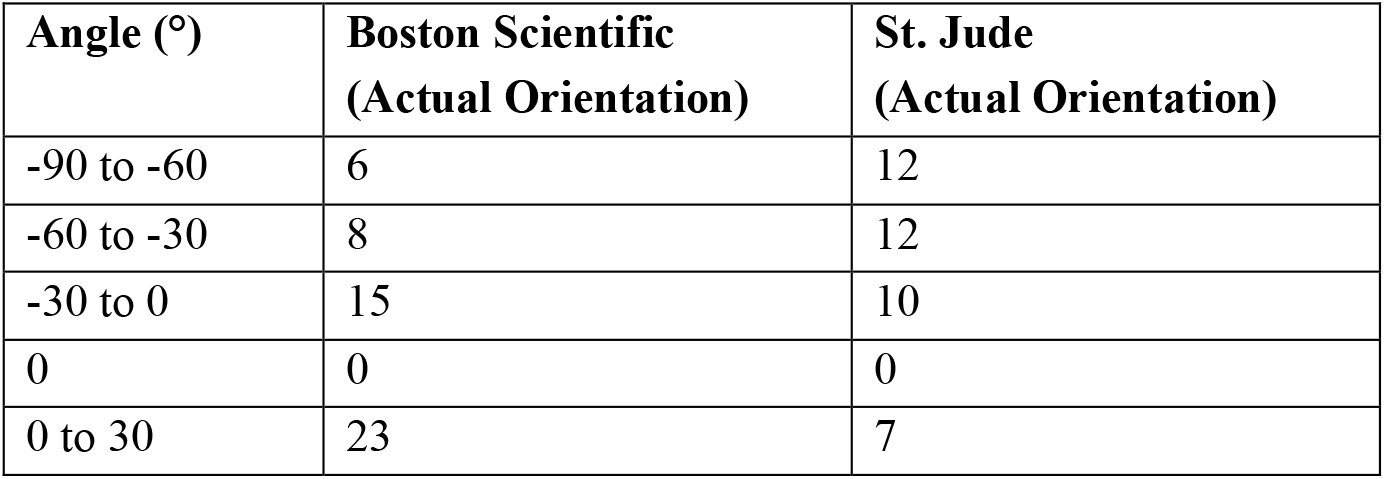
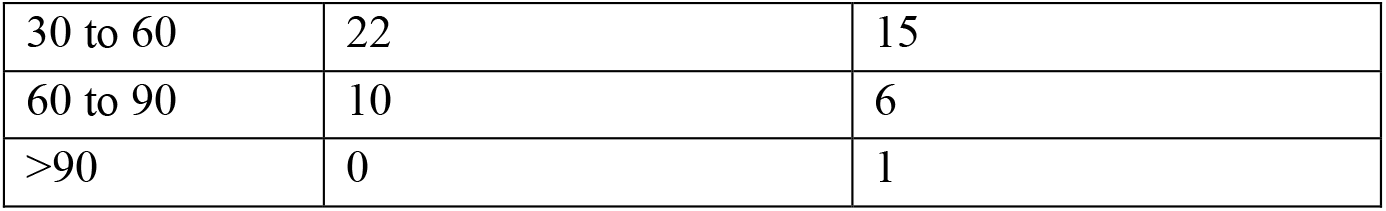
Distribution of the actual and intended lead orientations across both DBS systems and medical institutions, as calculated with DiODe.

### Deviation from intended orientation: Northwestern Memorial Hospital

The distributions of the deviation from the intended orientations for each DBS system are illustrated in Figure 4. Deviations from -89° to 73° with respect to the intended orientation were observed for Boston Scientific Cartesia™ leads, and from -79° to 92° for the St. Jude leads, respectively. Deviations of more than 30° occurred in 34 Boston Scientific leads (51.5%) and in 16 St. Jude leads (72.7%). Deviations of more than 60° occurred in 12 Boston Scientific leads (18.2%) and in 9 St. Jude leads (40.9%). The median deviation was 32° (IQR=39°) for Boston Scientific leads and did differ from the intended 0° orientation (p<0.001). The median deviation was 51° (IQR=41°) for St. Jude leads and did differ from the intended 0° orientation (p<0.001). There was not a significant bias between implanting towards the right or left for the St. Jude (p>0.05) leads, but the Boston Scientific leads were biased towards the left (p=0.014).

**Fig. 4.**
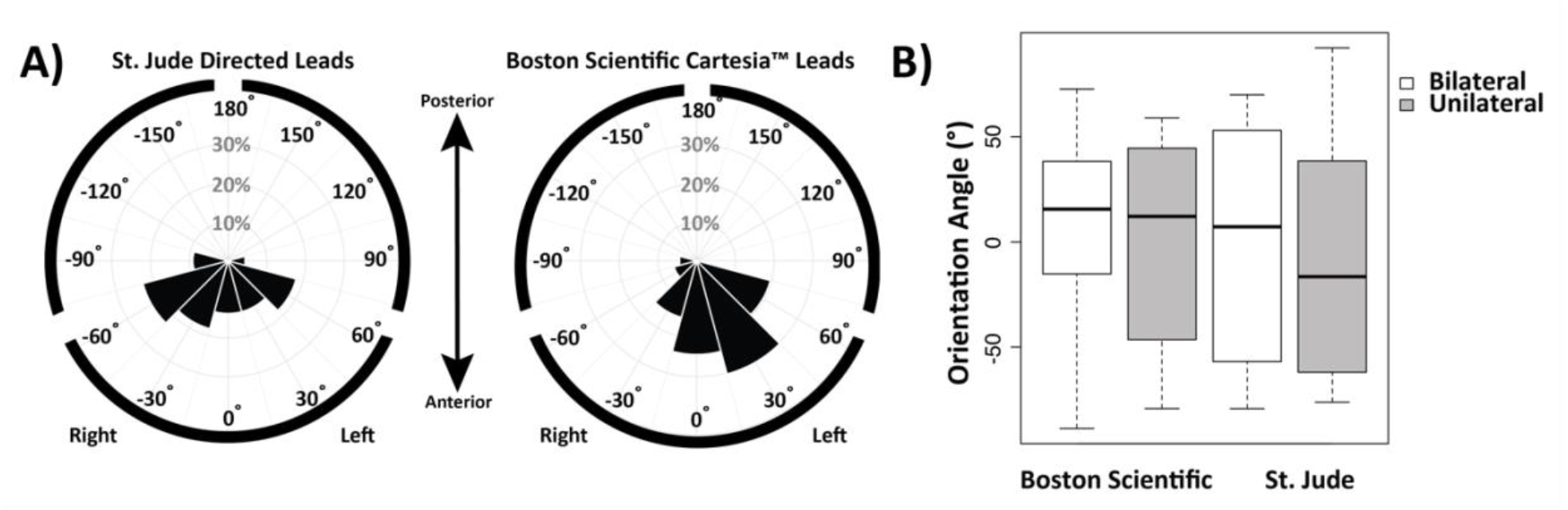
(A) Distributions of the actual orientation angles calculated with DiODe for leads intended to face anterior, showing only Northwestern Memorial Hospital patients, (B) Boxplot showing the distribution of the actual orientation angles for unilateral and bilateral patients separately.

Unilateral patients showed deviations from the intended 0° between -79° and 92°, while bilateral patients had deviations between -89° and 73°. The median deviation was -5° (IQR=45°) for the unilateral patients and 16° (IQR=38°) for the bilateral patients (Fig. 4B).

### Deviation from intended orientation: Albany Medical Center

The distributions of the deviation from the intended orientations for each DBS system are illustrated in Figure 5. Deviations from -64° to 86° with respect to the intended orientation were observed for Boston Scientific Cartesia™ leads, and from -88° to 81° for the St. Jude leads, respectively. Deviations of more than 30° occurred in 12 Boston Scientific leads (66.7%) and in 30 St. Jude leads (73.2%). Deviations of more than 60° occurred in 4 Boston Scientific leads (22.2%) and in 10 St. Jude leads (24.4%). The median deviation was 41° (IQR=42°) for Boston Scientific leads and did differ from the intended 0° orientation (p<0.001). The median deviation was 40° (IQR=32°) for St. Jude leads and did differ from the intended 0° orientation (p<0.001). There was not a significant bias between implanting towards the right or left for the St. Jude leads (p>0.05), but the Boston Scientific leads were biased towards the left (p=0.029). Unilateral patients showed deviations from the intended 0° between -63° and 77°, while bilateral patients had deviations between -88° and 86°. The median deviation was 32° (IQR=31°) for the unilateral patients and 6° (IQR=32°) for the bilateral patients (Fig. 5B).

**Fig. 5.**
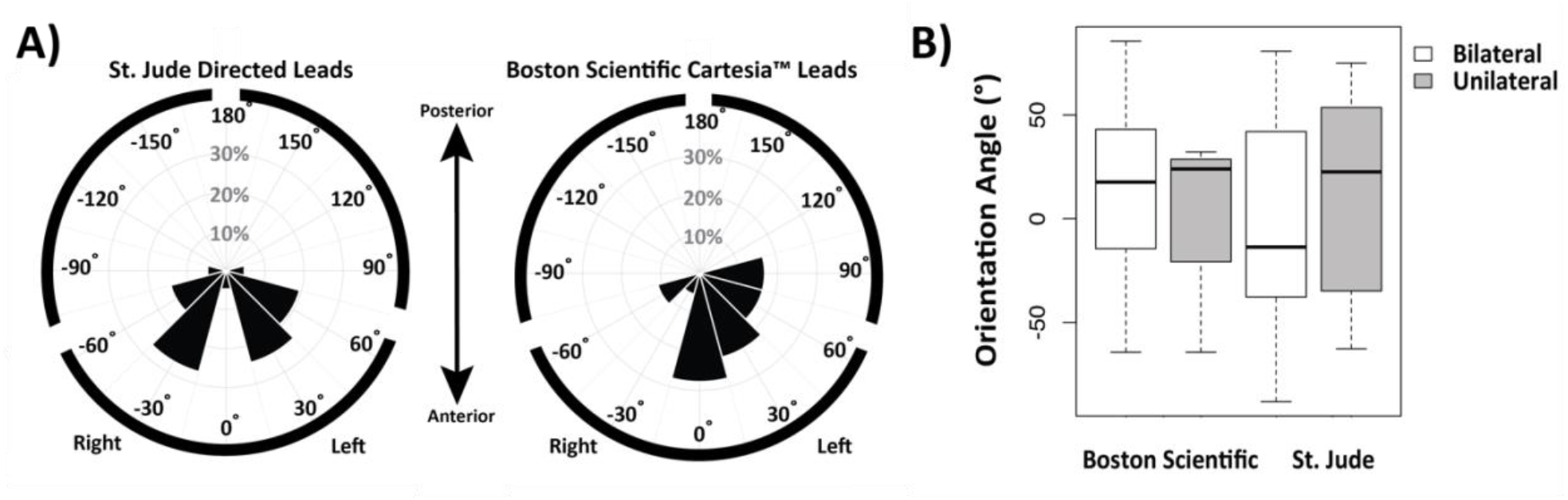
(A) Distributions of the actual orientation angles calculated with DiODe for leads intended to face anterior, showing only Albany Medical Center patients, (B) Boxplot showing the distribution of the actual orientation angles for unilateral and bilateral patients separately.

## Discussion

In our sample size of 104 patients, 57 patients with Boston Scientific Cartesia™ and 47 patients with St. Jude Directed leads from two institutions, Northwestern Memorial Hospital and Albany Medical Center, we discovered a statistically significant rotation that occurred across institutions, regardless of lead design. These results are in agreement with Dembek et al. (26) and Lange et al. (27), who have previously indicated that DBS leads deviate significantly from the surgically intended orientation. However, in contrast to previous studies that did not find a bias toward a certain direction, we have discovered that lead orientations were biased toward left for Boston Scientific leads across both institutions.

Additionally, we discovered that there was a large discrepancy in the orientation angles calculated by the three users using the manual refinement tool within the DiODe algorithm. Across both institutions, there was a 10° difference in the orientation angles calculated by at least two users in 30% of the Boston Scientific leads and 61% of the St. Jude leads. Understanding that a 30° change in orientation is considered significant, it is alarming that we still calculated at least a 30° difference in the orientation angles from at least two users in 12% of the Boston Scientific leads and 40% of the St. Jude leads. This large disagreement between users has not been acknowledged in previous studies, but may have impacted their results. We speculate that the larger discrepancy in values for St. Jude’s leads was partially due to the fact that DiODe has yet to be rigorously phantom validated for this particular lead design, despite the fact that St. Jude’s CT lead artifacts are similar to those created by the Boston Scientific leads.

Other factors may have affected our findings, such as the dates of implantations of each lead. When bilateral leads are not implanted on the same day, it is possible to see the shifting of the first lead due to torque during implantation or the fixation to the skull of the second lead (33). Only 2.04% of the patients analyzed in this study underwent the full bilateral implantation on the same day; therefore, the first lead may have been implanted facing the intended orientation and then later rotated during the second lead implantation. For each patient, we used the CT scan taken after the second lead implantation, so it is possible that the first implanted lead underwent an additional rotation.

Overall, our results agree with previous studies that have shown that there is a significant lead rotation from the intended surgical orientation, and additionally raises questions into the user error that occurs when manual refining the results calculated by DiODe. Further research is needed to evaluate why user discrepancies are so high and to establish a uniform process for all users to achieve the same (within 10°) orientation angle if using the manual refine tool in the algorithm. Additionally, we highly recommend that actual lead orientations are incorporated into lead modeling pipelines, especially those outside of Lead-DBS, and that all studies utilizing the DiODe toolbox should evaluate the orientation angles across multiple users until this discrepancy is reduced.

## Data Availability

All data produced in the present study are available upon reasonable request to the authors

